# Targeting fronto-limbic dysfunctions via intermittent theta burst stimulation as a novel treatment for Functional Movement Disorders

**DOI:** 10.1101/2021.02.01.21250621

**Authors:** Primavera A Spagnolo, Jacob Parker, Silvina Horovitz, Mark Hallett

**Author notes:** Correspondence should be addressed to: Primavera A Spagnolo, MD, PhD, Connors Center for Women’s Health and Gender Biology, AND Department of Psychiatry, Brigham and Women’s Hospital, Harvard Medical School, Thorn Building, 75 Francis St, Boston, MA 02115, Mark Hallett, MD, DM (hon), Human Motor Control Section, Medical Neurology Branch, National Institute of Neurological Disorders and Stroke, National Institutes of Health, Building 10, Room 7D37, 10 Center Drive, Bethesda, MD 20892-1428.

## Abstract

**Background:** Neuroimaging studies suggest that corticolimbic dysfunctions, including increased amygdala reactivity to emotional stimuli and heightened fronto-amygdala coupling, play a central role in the pathophysiology of functional movement disorders (FMD), although there is no direct causal evidence of this relationship. Transcranial magnetic stimulation (TMS) has the potential to probe and modulate brain networks implicated in neuropsychiatric disorders, including FMD. Therefore, in this proof-of-concept study, we investigated safety, tolerability and preliminary efficacy of fronto-amygdala neuromodulation via targeted left prefrontal intermittent theta burst stimulation (iTBS) on brain and behavioral manifestations of FMD.

**Methods:** Six subjects with a clinically defined diagnosis of FMD received three open-label iTBS sessions per day, for two consecutive study visits. Safety and tolerability were assessed throughout the trial. Amygdala reactivity to emotionally valenced stimuli presented during an fMRI task and fronto-amygdala connectivity at rest were evaluated at baseline and after each stimulation visit, together with subjective levels of arousal and valence in response to affective stimuli. FMD symptom severity was assessed at baseline, during treatment and 24 hours after receiving the last iTBS session.

**Results:** Multiples doses of iTBS were well-tolerated by all participants. Intermittent TBS significantly decreased fronto-amygdala connectivity and also influenced amygdala reactivity to emotional stimuli. These neurocircuitry changes were associated to a significant decrease in negative valence and an increase in positive valence levels following iTBS. Furthermore, we also observed a marked reduction in FMD symptom severity post stimulation.

**Conclusions:** Corticolimbic modulation via iTBS represents a promising treatment for FMD that warrants additional research.

## Introduction

Functional movement disorders (FMD), part of the wide spectrum of functional neurological disorders, represent one of the most common conditions encountered by neurologists and neuropsychiatrists today (1). Despite the high prevalence and the considerable individual and societal burden associated with these disorders, their etiopathogenesis is still elusive and only few evidence-based treatments are currently available for patients with FMD (2).

In recent years, however, there have been substantial advances in the pathophysiologic understanding of these disorders, which have critically impacted on the therapeutic management of FMD (3). In particular, functional neuroimaging studies have provided extensive evidence of alterations in activity and connectivity in brain networks mediating motor functions, cognitive control, emotion processing, and perceptual awareness (4,5). Among these alterations, cortico-limbic dysfunctions appear to play a central role in the pathophysiology of FMD. Indeed, increased amygdala reactivity to emotionally valenced stimuli has been repeatedly reported in patients with FMD, together with heightened amygdala coupling to motor and cognitive control regions, and abnormal activation of several prefrontal regions during emotion processing tasks (6–8). These findings suggest that these disorders are characterized by an impairment in bottom-up and top-down mechanisms in cortico-limbic circuits during emotion processing, which may exert an abnormal influence on motor systems (9,10).

This emerging disease model of FMD as a disorder of impaired neurocircuitry provides a robust rationale for the development of neurobiologically-based treatments aimed at targeting and remodeling dysregulated brain circuits. This hypothesis is supported by preliminary findings showing that the efficacy of motor rehabilitation and cognitive behavioral therapy in reducing functional motor symptoms is associated with changes in brain regions and networks implicated in FMD (11,12).

A further promising intervention garnering significant attention is transcranial magnetic stimulation (TMS). This non-invasive brain stimulation technique alters cortical excitability in the targeted cortex as well as in its downstream interconnections, via transcranial delivery of magnetic pulses at different intensities and frequencies. By applying a patterned, repetitive sequence of TMS (rTMS), temporally sustained changes in neuronal firing can be achieved, causing changes in brain functions and behavior (13). For instance, rTMS can affect cognitive processes, such as attentional control, as well as response to rewards and affective stimuli (14). These effects could be long-lasting following multiple stimulation sessions and are thought to reflect the ability of rTMS to enhance synaptic plasticity in a dose-dependent manner (15). This mechanism is thought to underlie the therapeutic effects of rTMS, which is currently approved by the Food and Drug Administration for the treatment of depression, OCD and nicotine addiction.

In the field of FMD, the use of rTMS as a therapeutic tool is still in its infancy. The published literature includes case report series and a small number of sham-controlled trials, characterized by a substantial variability in design and stimulation parameters (16–19). Importantly, these studies evaluated rTMS effects on clinical and behavioral outcomes of FMD, while it is unknown whether rTMS induces neuromodulatory and neuroplastic changes in relevant brain networks, which may underlie the observed clinical effects.

To begin addressing these questions, we conducted a pilot, proof-of-concept study in a sample of patients with hyperkinetic FMD with the aim to: i) assess the safety and feasibility of multiple sessions of an accelerated intermittent theta burst stimulation (iTBS) protocol (3600 pulses); and ii) uncover the effects of prefrontal iTBS on FMD-associated cortico-limbic dysfunctions (amygdala hyperreactivity to emotional stimuli and increased fronto-amygdala connectivity).

Targeting the left dorsolateral prefrontal cortex (DLPFC) via excitatory rTMS and iTBS has been shown to improve emotion processing in both healthy and patient populations, by inhibiting negative bias and increasing response for positive stimuli (20–22), while right-sided prefrontal stimulation is associated with increased right amygdala activation and enhanced attentional allocation to threatening stimuli (23,24). Furthermore, it has been reported that left prefrontal iTBS also decreases functional connectivity between DLPFC and limbic regions (25,26), which is increased in individuals with FMD (8).

On the bases of these observations, we hypothesized that an accelerated protocol of left DLPFC iTBS in individuals with FMD would normalize fronto-amygdala hyperconnectivity and promote a dissociative pattern of amygdala response to negative and positive emotional stimuli presented during functional magnetic resonance imaging (fMRI) sessions. To obtain further insights into the mechanisms of action of iTBS, we also explored the effects of stimulation on subjective valence and arousal levels in response to affective stimuli, and on FMD symptomatology.

## Materials and Methods

### Participants

Six patients with clinically definite FMD, as diagnosed by at least two movement disorder specialists using Fahn and Williams criteria (27), were recruited from the Human Motor Control Clinic at the NIH between February 2018 and May 2019. Participants belonged to a larger ongoing study investigating the clinical and neurobiological correlates of FMD. Exclusion criteria included comorbid neurologic diseases; lifetime history of psychosis or bipolar disorder, current diagnosis of alcohol and substance use disorders, obsessive-compulsive disorder, major depressive disorder or post-traumatic stress disorder; current suicidality; any change in psychoactive pharmacotherapy in the 4 weeks prior to the study. Subjects were also excluded if they had contraindications to TMS administration and/or MRI, including therapy with pro-convulsant medications, history of traumatic brain injury; any personal or family history (1st degree relatives) of seizures other than febrile childhood seizures; abnormal clinical MRI brain; hearing loss; tinnitus and pregnancy/lactation.

We performed the experiments in agreement with relevant guidelines and regulations (28,29). The NIH Institutional Review Board approved the study. All participants provided written informed consent.

### Study design

This was a proof-of-concept, feasibility, open-label study which included four outpatient visits (Figure 1). On visit 1 (V1), we evaluated study eligibility, obtained informed consent and performed a comprehensive assessment of the clinical phenotype of study subjects, through neurological exam, the semi-structured clinical interview for psychiatric diagnosis (SCID-IV-R) (30) and a battery of self-report and clinician-rated scales and questionnaires [for details see *Supplemental Material*]. Further, participants underwent a baseline MRI scanning session to acquire T1-weighted, resting-state functional connectivity (rsFC) and task-based data. In addition to evaluating DLPFC-amygdala FC prior to the stimulation, the rsFC data were used to identify the cortical iTBS target within the left DLPFC that was functionally connected to the left amygdala, as described below.

**Figure 1.**
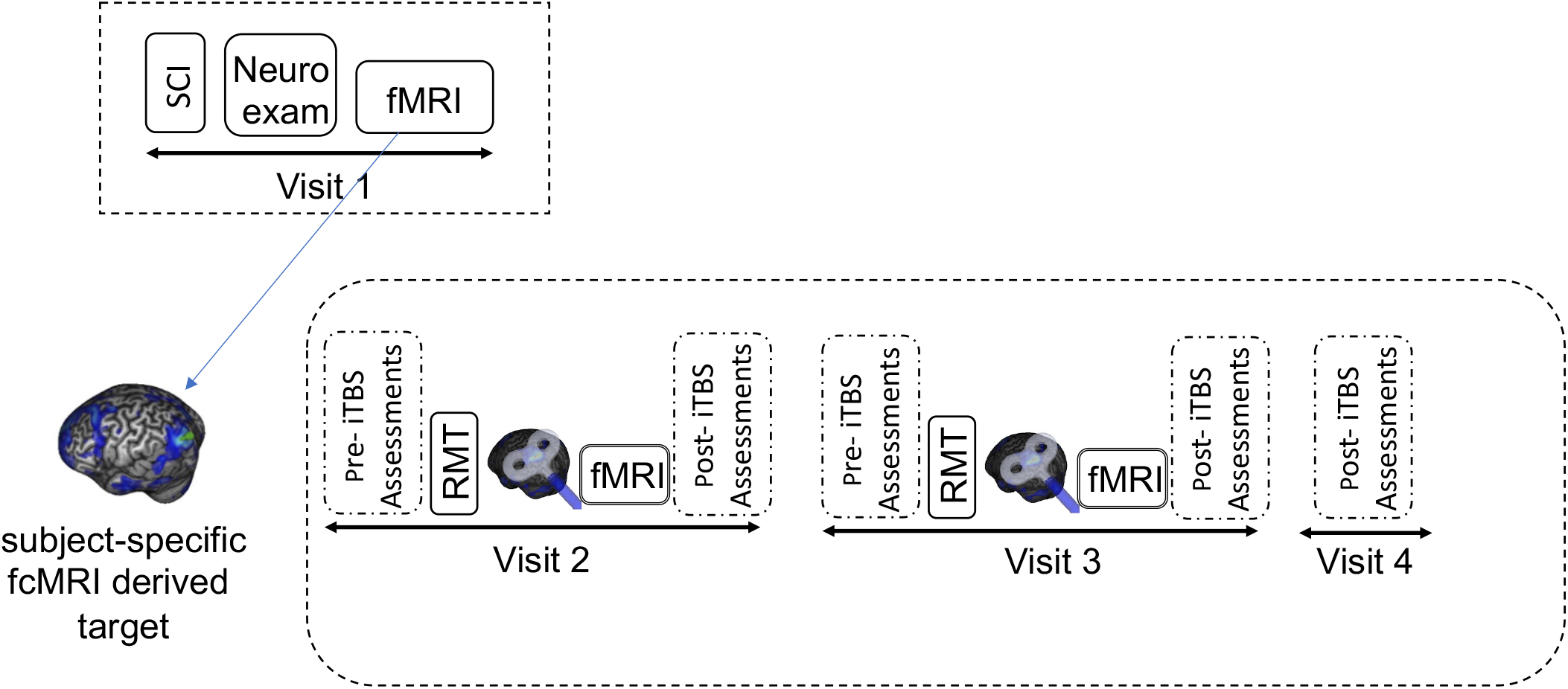
Schematic representation of the study design. During Visit 1, we obtained the informed consent and conducted a structured clinical interview [SCI], which included several patient- and clinician-rated scales and questionnaires (see main text and Supplemental Material for details). We also conducted a neurological examination and assessed FMD symptom severity using the Simplified-Functional Movement Disorder Rating Scale (S-FMDRS). After this, we acquired structural and functional (fMRI) images. Personalized targets were identified using each subject’s resting-state fMRI data. One week after Visit 1, we carried out three consecutive visits. Visit 2 and 3: during each of this visit, participants underwent pre-iTBS assessments, which included completing the S-FMDRS and the TMS and MRI safety screening forms. We then determined resting motor threshold (RMT) using single pulse TMS and administered three sessions of iTBS (1800 pulses/daily), followed by an fMRI scan. Subjects also completed several post-iTBS assessments, which included the S-FMDRS, the Positive and Negative Affect Scale (PANAS) and Young Mania Rating Scale (YMRS) as well as the iTBS side effects monitoring form. Visit 4 was a follow-up visit during which we repeated the post-iTBS assessments.

Following the MRI session, we measured subjective levels of arousal and valence in response to affective face stimuli, using the self-assessment Manikin (SAM) (31). The SAM is a pictorial rating scale that directly measure the affective dimension of valence and arousal in response to emotionally valenced stimuli.

After a period of 1-2 weeks from visit 1, subjects participated in three consecutive outpatient visits, which were separated by a 24-hr interval. During visit 2 and 3 (V2 and V3), they received a block of three sessions of iTBS to the individualized left DLPFC target mapped using an online MRI-guided neuronavigation system. Approximately 15-20 minutes after the last iTBS session, participants underwent an fMRI scan (task-based and rsfMRI), and then provided valence and arousal levels in response to emotional stimuli, as described above.

We also collected several additional clinical and subjective measures, including severity and frequency of functional motor symptoms, which was assessed at 6 different timepoints (baseline, V2: pre- and post-iTBS; V3: pre- and post-iTBS, and at V4) using a modified version of the Simplified-Functional Movement Disorders Rating Scale (S-FMDRS) (32). Furthermore, following each stimulation block, we administered the iTBS monitoring questionnaire, an internally generated 13-item interview-based yes/no questionnaire assessing side effects of iTBS (e.g., headaches, nausea, seizure), and the Positive and Negative Affect Scale (33) and Young Mania Rating Scale (34), to evaluate potential iTBS-induced changes in mood.

The last outpatient visit (V4) was a follow-up visit, during which iTBS-related side effects and FMD symptoms were again assessed 24 hours after receiving the last stimulation session.

### TMS and iTBS protocol

We delivered single pulse TMS and iTBS using a Magstim Rapid machine (Magstim Co., Whitland, South West Wales, UK) and a “figure of 8” cooled butterfly coil. Single pulse TMS was used to localize the motor hotspot and determine the resting motor threshold (RMT), using the adaptive threshold hunting procedure (35). We used an accelerated iTBS protocol, which entailed three iTBS sessions per day, with at least a 20-min interval between sessions. The duration of the inter-session interval was based on previous studies indicating that a period ≥15 minutes between consecutive iTBS blocks enhances cortical plasticity (36). Further details on stimulation parameters, neuronavigation procedures and context of stimulation are provided in the *Supplemental Material*.

### Image Acquisition

Imaging was acquired during each visit with a 3-T MR750 GE scanner using a 32-channel head coil. Each fMRI scan included five consecutive runs in the following order: anatomical scan (∼ 5 min); resting state (∼ 6 min), task (2 runs, ∼ 10 min each), and resting state (∼ 5 min). The full parameters are provided in the *Supplemental Material*.

### fMRI Task and Stimuli

Processing of emotional faces was measured using a modified version of the procedures described in Voon et al. (9). Subjects viewed affective face stimuli in three different block conditions of fearful, happy and neutral stimuli. Further details are provided in the *Supplemental Material*.

### Imaging processing and data analysis

The fMRI data from each subject were processed using AFNI (v16.2.16; (37) afni.nimh.nih.gov). Resting-state FC data was analyzed using the first (pre-task) run of each study visit. The left amygdala and the individual DLPFC iTBS targets were defined as regions of interest (ROIs). Pearson’s *r* was computed between the average BOLD time series of the left amygdala, as defined in cytoarchitectonic \ [(38); −23, −4, −20; 63 voxels 3×3×3) and the individual DLPFC stimulation target, which was derived using a 6-mm radius sphere region of interest (ROI) centered on the coordinates of the iTBS targets (see section below). Then, we applied *r*-to-*z* Fisher transformation and tested whether multiple sessions (‘doses’) of iTBS induced changes in left amygdala - DLPFC target, using a mixed linear effects model. The FC between the left Amygdala and the whole brain was computed in a similar manner and used as a covariate to control for individual differences in whole brain FC. Given that the left and right amygdala are highly connected, we also explored whether iTBS also exert a neuromodulatory effect on right amygdala - DLPFC target FC.

For the task-fMRI data, we performed a ROI-based analysis, with the left amygdala selected as the ROI, and compared the fear-happy linear contrast over study visits, using a mixed linear effects model. We repeated this analysis also to explore iTBS-induced changes in right amygdala activation patterns.

A complete description of the image preprocessing and analysis procedures, including ROI definition, is provided in the *Supplemental Material*.

### Target Identification

To identify the individualized iTBS target in the left DLPFC, we used rsFC data (pre-task run) collected at V1. Pearson’s *r* was computed between the BOLD time series of each voxel in the left DLPFC and the average BOLD time series of the left amygdala. The coordinates of the voxel with the highest *r* value in the left DLPFC was identified as the target of iTBS, which was then marked on the individual structural MRI using the neuronavigation software Brainsight, Rogue Research, Inc., United Kingdom).

This connectivity-based targeting approach offers the advantage to generate individualized stimulation sites, and also allows to target remote, but functionally interconnected, brain regions. This could be particularly relevant considering that the average clinical efficacy of left DLPFC TMS is related to intrinsic functional connectivity between the TMS target and distal brain regions (39).

### Statistical Analysis

Mixed linear effects models were used to test the effects of multiple doses of iTBS on the imaging, behavioral and clinical outcome measures collected in this study. A detailed description of data analyses is provided in the *Supplemental Material*. All statistical analyses were performed using R V.3.0.2. The level of significance for all tests was set to 0.05.

## Results

Study subjects (3 males and 3 females; ages 24–57 y, mean = 48.8, SD = 12.3) reported an average disease duration of 10.7 years (± 7), with a baseline S-FMDRS score of 14.5 (± 6.8). The abnormal movements reported included tremor and jerking movements (75%), abnormal gait/balance (65%), abnormal speech (50%), abnormal posturing/dystonia (35%). Four subjects had a lifetime history of depressive and/or anxiety disorders and only one subject had a current diagnosis of generalized anxiety disorder. Both depressive and anxiety symptom severity were in the mild range (Hamilton Depression Scale= 2.7 ± 1; Hamilton Anxiety Scale =8 ± 9.1).

Due to motion artifacts, baseline rsFC data were not available for one subject, who was excluded from the analysis (V1-V3: *n*= 5). Task-based data were collected at all timepoints except for one subject, who could not perform the fMRI task on V3 for an unexpected scanner shutdown (V1: *n* = 6; V2: *n* = 6; V3: *n* = 5).

### Safety and Tolerability

All participants completed the entire study protocol, without unexpected, serious adverse events. Four subjects reported mild, transient (< 60 minutes) headache, beginning during or shortly after the first iTBS session (Visit 2). Three subjects experienced headache also following the second stimulation visit (Visit 3). Headaches resolved without intervention, with the exception of one participant who required a single dose of acetaminophen. Two participants reported mild to moderate scalp pain after the first iTBS block. No negative side-effects in affective assessments were reported or observed after iTBS. No participant experienced any signs of mania or suicidality. The iTBS Monitoring Questionnaire revealed no seizures, fainting, difficulties speaking or understanding speech, or impairment of thought.

### Functional MRI

#### Left Amygdala – DLPFC iTBS target resting-state functional connectivity

After two blocks of iTBS (3 daily sessions; 3600 pulses), we observed a significant iTBS dose effect on the functional connectivity between the left amygdala and the individualized iTBS DLPFC (*t* = - 2.404, *p* = 0.047), which decreased from baseline (V1) to V3, as shown in Figure 2a. Exploratory analysis using the right amygdala as the ROI also found a significant effect of dose on connectivity between this region and the iTBS target (*t* = -3.095, *p* = 0.018) [Suppl. Fig. 1a].

**Figure 2.**
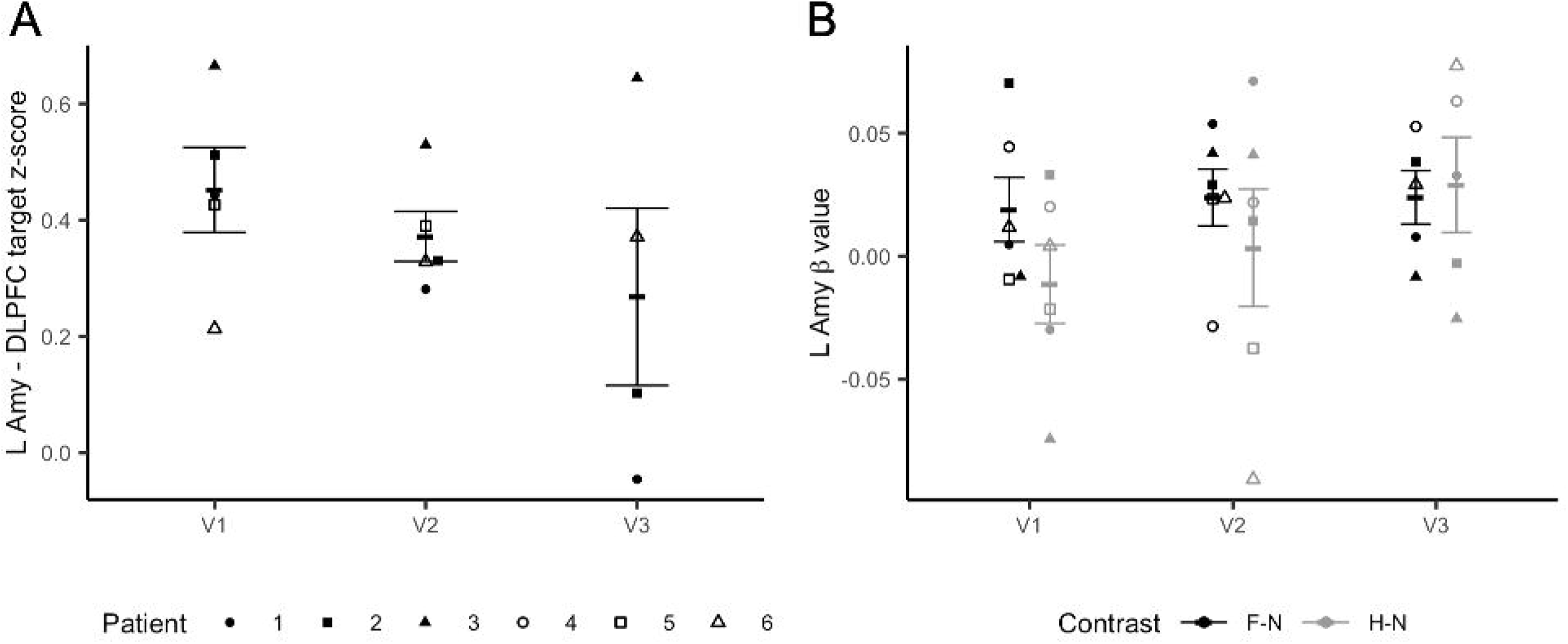
Imaging results. Panel A shows changes in resting state functional connectivity (rsFC) between the individualized dorsolateral prefrontal cortex (DLPFC) target and the left amygdala. The y axis represents changes in z-transformed region-to-region correlation strength as a result of iTBS. Panel B shows changes in left amygdala reactivity to fearful to neutral (F-N) vs. happy – neutral (H-N) faces during the fMRI task. The y axis represents left amygdala beta values. Error bars represent SEM.

#### Left Amygdala Reactivity

Linear mixed models found a trend level dose effect of iTBS on left amygdala activation in response to positive-negative emotional stimuli (happy – fearful facial stimuli) (*t* = -2.182, *p* = 0.057). Specifically, at baseline (V1) there was a difference in left amygdala response to happy vs fearful faces (*t* = -3.518, *p* = 0.017). This difference was not significant after iTBS (V2: *t* = -0.852, *p* = 0.43; V3: *t* = 0.323, *p* = 0.76). The changes can be explained by modulations to the response to happy faces. The plot in Figure 2b shows that while neural response to fearful vs. neutral faces remained substantially stable after both iTBS sessions, left amygdala reactivity to happy vs. neutral faces increased after receiving both iTBS blocks. Conversely, we did not find an iTBS dose effect on right amygdala response to fearful-happy faces (*t* = -1.846; *p* = 0.0979) [Suppl. Fig. 1b].

### Valence and Arousal Levels

Consistently with the imaging results, we observed a significant iTBS dose effect on valence ratings associated to fearful faces (*t* = -3.069, *p* = 0.012), which decreased at V3 when compared to baseline (V1-V3: *t* = -2.912, *p* = 0.017) [Fig. 3a]. These changes were accompanied by a marked dose effect on positive valence ratings (*t* = 10.784, *p* <.0001), which increased at V3 compared to baseline (V1-V3: *t* = 11.276, *p* <.0001) [Fig. 3b]. No effects of iTBS on arousal levels in response to both positive and negative emotional stimuli were found (V1-V3: *t* = - 1.738, *p* = 0.116) [*data not shown*].

**Figure 3.**
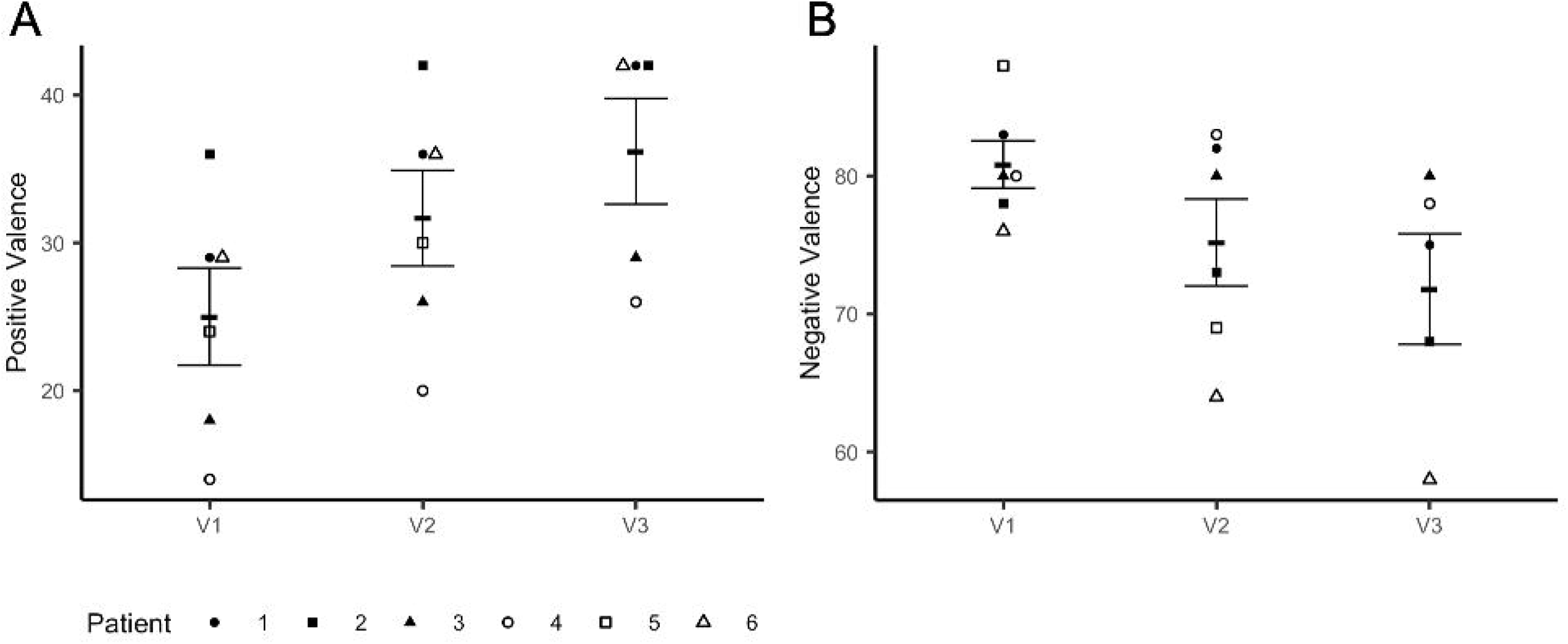
Changes in Positive Valence (A) and Negative Valence (B) levels.

### Simplified-Functional Movement Disorder Rating Scale Scores

Linear mixed-model analysis revealed a significant decrease in FMD symptom severity from pre-to post-iTBS across both stimulation visits, as measured by S-FMDRS scores (t = 5.439, *p* <.0001) [Fig. 4]. This decrease was significant within each stimulation visit (pre-to post-iTBS V2: *t* = 4.715, *p* = 0.0053; pre-to post-iTBS V3: *t* = 3.266, *p* = 0.0309) and between pre-iTBS scores collected at V2 and those at the follow-up visit (V4) (*t* = -5.490, *p* = 0.0004). There was no difference between S-FMDRS scores collected at V1 (baseline) and those measured at V2 prior to the stimulation session (: *t* = 0.498, *p* = 0.640).

**Figure 4.**
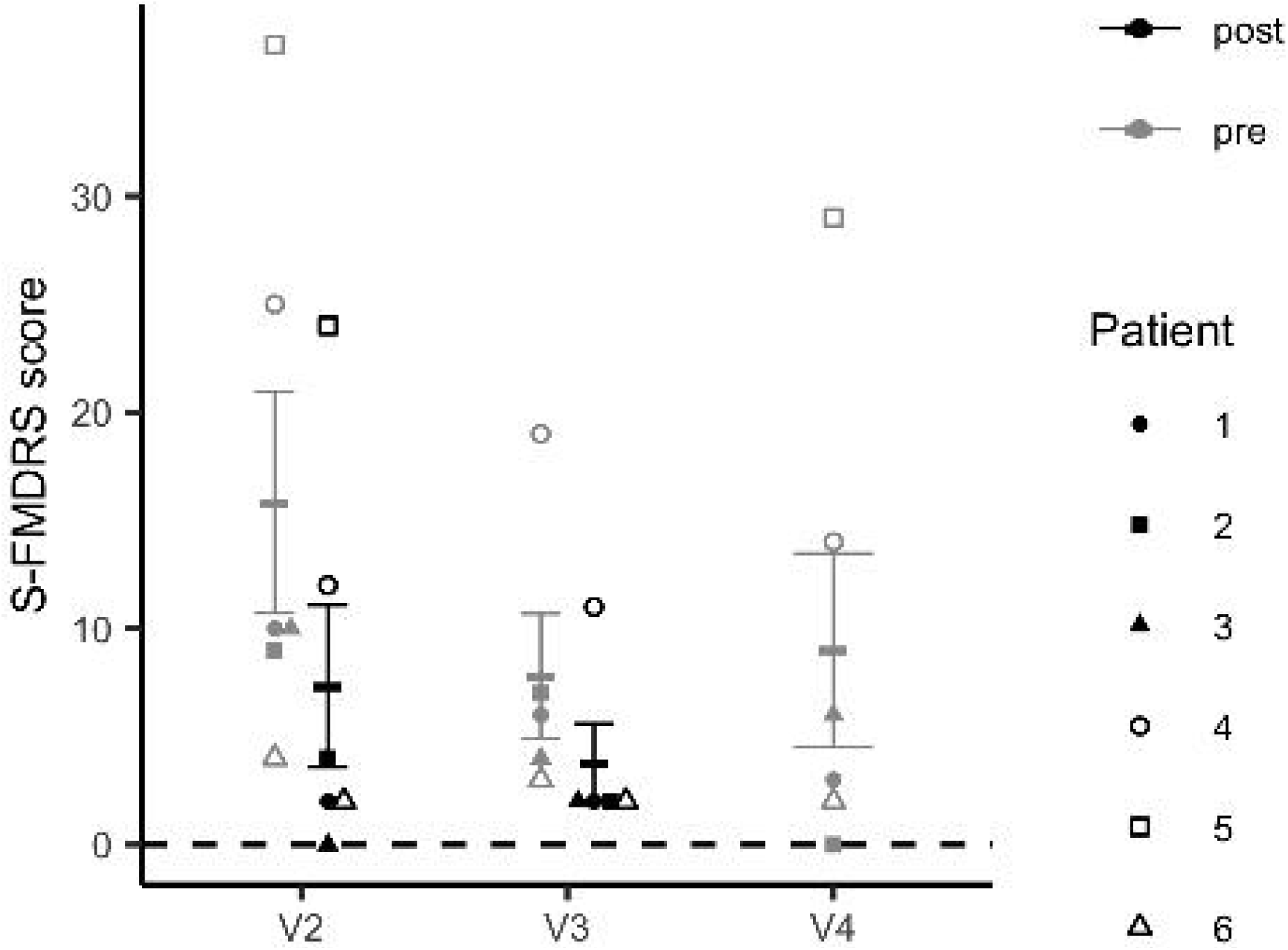
Changes in Simplified-Functional Movement Disorder Rating Scale (S-FMDRS) scores.

## Discussion

This study is to our knowledge the first investigating the effects of left prefrontal iTBS on neurocircuitry, behavioral and clinical manifestations of FMD. First, we showed that an accelerated iTBS protocol was well-tolerated with a good safety profile. The most frequently reported side effect was occasional mild headache, which remitted either spontaneously or following acetaminophen administration. Second, we found that multiples doses of iTBS targeting functionally relevant sites within the left DLPFC decreased fronto-amygdala connectivity and also influenced amygdala reactivity to emotional stimuli during an affective face perception task. These neurocircuitry changes were associated to variations in subjective indices of emotion processing, with valence levels in response to negative stimuli decreasing following iTBS. Furthermore, we also observed a significant reduction in FMD symptom severity post stimulation, as measured by the S-FMDRS.

Our finding of decreased corticolimbic rsFC after iTBS parallel results from previous studies showing that targeted prefrontal stimulation reduced both within- and between-networks connectivity (40). Specifically, active iTBS compared to sham has been shown to decrease the connectivity between DLPFC and insula (26), and between the DLPFC and nodes of the default mode network (25), both in healthy volunteers. Importantly, recent research has suggested that iTBS efficacy in patients with several psychiatric disorders may depend upon ‘normalization’ of pathologically increased connectivity (41). This mechanism may also contribute to the effects of prefrontal stimulation in FMD patients. Morris and colleagues (8) reported that individuals with FMD compared to controls exhibit elevated rsFC between amygdala and DLPFC, in line with the direction of connectivity observed at baseline in our study sample. This abnormal connectivity pattern differentiates FMD patients from those with mood and anxiety disorders, who generally show reduced connectivity between these regions. Therefore, heightened DLPFC-amygdala coupling may represent a specific brain feature of FMD, which can be targeted and remodeled via iTBS.

In addition to the effects on corticolimbic rsFC, we also observed a marginal effect of prefrontal iTBS on amygdala reactivity to affective face stimuli. Specifically, during V1 (baseline) left amygdala activation to fearful vs. neutral faces significantly differed from happy vs. neutral faces. This finding is consistent with a prior study reporting enhanced left amygdala activation to negative vs. neutral stimuli in FMD patients (10). Importantly, the authors found that the magnitude of amygdala response elicited by negative stimuli increased following repeated exposure. Impaired amygdala habituation to emotional stimuli has been repeatedly reported in individuals with FMD (6). In our study, targeted prefrontal iTBS appears to prevent amygdala sensitization to negative stimuli, while increasing response to positive stimuli. Confirmatory behavioral analyses indicated that changes in amygdala reactivity were accompanied by a decrease in negative valence and an increase in positive valence ratings following multiple doses of iTBS, which may reflect changes in the salience attributed to affective stimuli. Together, these results parallel previous studies showing that left DLPFC stimulation improves emotion processing by inhibiting negative bias and improving affective processing of positive stimuli (14,20–23).

Conversely, our exploratory analysis did not reveal an effect of iTBS on right amygdala activation during the affective face perception task. This lack of effects could be due to the different roles that left and right amygdalae appear to play in emotion processing, as suggested by neuroimaging studies (42). Studies in patients with FMD have reported alterations in both right and left amygdala during emotion processing, with the right amygdala showing heightened reactivity to emotional stimuli, regardless of their valence (9), and the left amygdala exhibiting both hyperactivity and sensitization to negative stimuli (10). In the current study, we chose to modulate the left prefrontal-amygdala circuitry given that this circuitry is directly implicated in processing aversive stimuli (43). Such stimuli have been shown to elicit defensive behavior and modulate motor function in FMD patients (44). Furthermore, right but not left DLPFC stimulation increases both right amygdala response to negative stimuli as well as anxious arousal levels (23,45), which could in turn trigger and/or exacerbate functional motor symptoms.

This hypothesis is in part supported by our observation that multiple doses of iTBS induced a marked reduction in FMD symptom severity, thus suggesting a direct causal role of fronto-amygdala circuitry in FMD, as indicated by an increasing body of neuroimaging studies (6). However, it should be noted that we did not compare the effects of left vs. right DLPFC stimulation, so future work is needed to test the laterality of this effect.

Our study should be interpreted in light of some limitations. First, this was an open-label study. All participants knew they would receive active iTBS, posing the risk for a placebo effect, as for any other intervention. However, we measured the effects of left prefrontal iTBS on brain, behavioral and clinical outcomes and found an effect of iTBS on both subjective and objective measures. Furthermore, the direction of the changes in fronto-amygdala rsFC and in amygdala activity during emotion processing was in line with previous sham-controlled TMS studies. Other limitations include a small sample size and lack of long-term follow-up assessment, which prevent from drawing strong conclusions.

As proof-of-concept, our study was conducted primarily to establish safety and feasibility of an accelerated iTBS protocol in patients with FMD, as well as of our entire study procedures (e.g., connectivity-based target selection, offline fMRI-iTBS paradigm). In addition, we also gained preliminary evidence that the prefrontal-amygdala circuitry may represent an effective target for neurocircuitry-based interventions for FMD. Larger sham-controlled, randomized clinical trials will be required to fully investigate the efficacy of this intervention, also in combination with other therapies for FMD, and to further characterize its mechanisms of action.

## Data Availability

The data that support the findings of this study are available from the corresponding author upon reasonable request.

## Contributorship

PAS and MH conceived the idea for this study and PAS wrote the manuscript. PAS and JP conducted the experiment; JP and SH performed the analysis of the imaging data. JP performed the statistical analysis. All authors provided critical feedback and helped shape the research, analysis and manuscript.

## Acknowledgments

This work was supported by NINDS intramural research program. The authors would like to thank Dr. Lisanby for her contribution in designing this study; Drs. Kathrin LaFaver and Dr. Carine Maurer for their help with patient recruitment and data collection; Patrick McGurrin for his help in conducting TMS procedures; and Elaine Considine for providing monitoring during study procedures. We also acknowledge the support of the clinical research staff in the NINDS intramural program.

## Full disclosures

M.H. is an inventor of patents held by NIH for an immunotoxin for the treatment of focal movement disorders and the H-coil for magnetic stimulation; in relation to the latter, he has received license fee payments from the NIH (from Brainsway). He is on the Medical Advisory Boards of CALA Health and Brainsway (both unpaid positions). He is on the Editorial Board of approximately 15 journals and receives royalties and/or honoraria from publishing from Cambridge University Press, Oxford University Press, Springer, and Elsevier. He has research grants from Medtronic, Inc. for a study of DBS for dystonia and CALA Health for studies of a device to suppress tremor.

