## Supplemental Materials for "Targeting fronto-limbic dysfunctions via intermittent theta burst stimulation as a novel treatment for Functional Movement Disorders"

**Supplemental Material**

*Exclusion/inclusion Criteria*

Exclusion criteria included comorbid neurologic diseases; lifetime history of psychosis or bipolar disorder, current diagnosis of alcohol and substance use disorders, obsessive-compulsive disorder, major depressive disorder or post-traumatic stress disorder; current suicidality; any change in psychoactive pharmacotherapy in the 4 weeks prior to the study. Subjects were also excluded if they had contraindications to TMS administration and/or MRI, including therapy with pro-convulsant medications, history of traumatic brain injury; any personal or family history (1st degree relatives) of seizures other than febrile childhood seizures; abnormal clinical MRI brain; hearing loss; tinnitus and pregnancy/lactation.

*Visit 1 – clinical and behavioral assessments*

During Visit 1, all participants were screened for psychiatric diagnoses using the Structured Clinical Interview for the Diagnostic and Statistical Manual of Mental Disorders, Edition IV, Text Revised (DSM-IV-R), Patient Edition (1). Anxious and depressive symptomatology were assessed using the Hamilton Anxiety Rating Scale (HAM-A) (2) and the Hamilton Rating Scale for Depression (HAM-D) (3). Participants also completed the Profile of Mood State (POMS) (4), the Young Mania Rating Scale (YMRS) (5), the Columbia Suicide Severity Rating Scale (C‐SSRS) (6) and the Symptoms Checklist 90-revised (SCL 90-R) (7). The Childhood Trauma Questionnaire (CTQ) (8) and the Traumatic Life Events Questionnaire (TLEQ) (9) were administered to assess exposure to stressful events during early life and adulthood. To establish contraindications to MRI and TMS administration, we administered two internally generated tools: the MRI safety screen questionnaire and the TMS safety screen questionnaire.

*MRI Procedures*

*Image Acquisition.* Imaging was acquired during each visit with a 3-T MR750 GE scanner using a 32-channel head coil. Structural images Structural scans were collected using a T1-weighted anatomical MRI (multi-echo magnetization-prepared rapid gradient echo [MEMPRAGE], voxel size 1 × 1 × 1 mm; repetition time [TR] 400 ms; echo time [TE] 1.69 ms; echo spacing 9.8 ms; number of echoes 4; bandwidth 650 Hz/Px; inversion time 1,100 ms; flip angle 7°; acceleration factor 2; matrix size 176 × 256 × 256; field of view [FoV] 256 mm, acquisition time: 6 minutes 2 seconds), while functional scans were acquired using a T2-weighted EPI sequence (voxel size 3 x 3 x 3 mm; number of slices 36; matrix size 36 x 72 x 72; TR 2500 ms; number of TRs 144 (resting state)/240 (task); TE 14.5 ms; echo spacing 17.8 ms; number of echoes 3). During resting state runs, participants were instructed to lie as still as possible in the scanner with their eyes closed, not to think about anything in particular and not fall asleep.

*fMRI Task and Stimuli.* Stimuli from the Karolinska Directed Emotional Faces (70 different faces; 35 male and 35 female posing happy, fearful, and neutral expressions, for a total of 210 stimuli) were used for the facial emotion processing task. The neutral face consisted of 25% happy morphed with 75% neutral as 100% neutral faces have previously been found to be aversive. Seventeen 24 s blocks (16 stimuli per block) of two runs were shown interspersed with 11 s fixation rest. Stimuli were presented centrally for 1 s with a fixation cross present for 0.5 s between trials. The images were randomized within blocks. All stimuli were back-projected onto a translucent screen behind the bore of the magnet, visible via an angled mirror placed above the participant’s head. To ensure that subjects attended the images, gender judgment button responses were acquired during the stimulus presentation period via a fiber optic button box. The facial emotion processing task is an implicit processing task; behavioral responses were not of interest and thus not included in the analyses. The task was performed using Presentation® software (Version 14.0, Neurobehavioral Systems, Inc., Berkeley, CA).

*Image Preprocessing.* Image preprocessing and analysis was completed using AFNI software (12). The MPRAGE for each visit was skull stripped and nonlinearly registered to the MNI ICBM 152 2009 template using the @SSwarper function. Each functional sequence was preprocessed using afni_proc.py with the following processing steps: despiking, slice-time correction, alignment to the processed MPRAGE*, registration to the MNI template space using the warps computed by @SSwarper*, co-registration of each functional volume*, multi-echo independent component analysis (via tedana.py), convolution with a 4-mm full-width at half-maximum Gaussian kernel, scaling each voxel time series to a mean of 100, and regression out of motion parameters. All spatial transformations (*) were concatenated and applied in a single step.

Volumes with excess motion (frame-wise displacement > 0.3mm) and/or with BOLD signal outliers in many voxels (proportion of voxels > 0.1) were censored.

*Region of Interests*. We selected three regions of interest (ROI): left amygdala, right amygdala and the individualized iTBS sites within the left DLPFC. For the right and left amygdala, ROIs were defined using a cytoarchitectonic atlas (13). For each amygdala ROI, the probability maps of the amygdala subdivisions were added together to form a probability map of the whole amygdala, which was threshold to a value of 0.5 to form the ROI. The DLPFC ROI was defined using a functional atlas that identified regions based on whole-brain resting-state functional connectivity (14).

*Task data analysis.* In order to compare task-related activation across conditions (happy, neutral, fearful), multiple linear regression was performed on the preprocessed task data using 3dDeconvolve within afni_proc.py. Regressors for each condition were created via convolution of the stimulus onset time series with the canonical hemodynamic response function. Stimuli were considered instantaneous for this analysis. Regression yielded condition-specific β-coefficient maps, which indicated task-related activation within each voxel. Linear contrast (Δβ) maps, which indicated the difference in activation between each pair of conditions within each voxel, were also computed.

*Intermittent TBS Procedures*

*Individualized iTBS Target Neuronavigation.* In order to allow for the use of standardized coordinates to mark the personalized iTBS target on the individual structural MRI within the Brainsight neuronavigation software, the processed MPRAGE was linearly registered to the MNI ICBM 152 2009 template. A linear transformation was used to conserve individual-specific spatial features, which were necessary to perform accurate landmark-based calibration of the coil prior to the iTBS session. This transformation was applied to the processed resting state sequence. Neuronavigation (Brainsight, Rogue Research, Inc., United Kingdom) with an optical tracking system (“Polaris Vicra”, Nothern Digital Inc., Canada) was used to ensure precise positioning of the coil over the individualized stimulation target within each session, across sessions, and across participants.

*Stimulation Parameters.* Each iTBS session consisted of 600 pulses in 50 Hz bursts of three pulses, separated by 200 ms (i.e., a 5 Hz frequency) for 2 s, followed by 8 s of no pulses over about 190 s (Huang et al., 2005). The magnetic field intensity for each session was set at 120% of that participant’s RMT and was increase/decrease in a ramp-like fashion at the onset and offset of each iTBS session, to minimize scalp discomfort/pain.

*Context of stimulation*. While receiving iTBS, participants wore earplugs and were seated in a comfortable chair, while watching a video extracted from a nature documentary on a 15-inch computer screen placed at a distance of 60 centimeters. This procedure allowed for a standardized ‘context of stimulation’ between sessions and across subjects.

*Data analysis*

*Resting-state functional connectivity (rsFC) data.* The effect of dose, or treatment accumulation over consecutive days, on the rsFC between the amygdala and the individualized iTBS target within the left DLPFC was tested using a mixed linear effects model. In this design, the rsFC was the outcome variable, dose was the continuous covariate of interest, the amygdala-whole brain rsFC was an additional covariate, and patient was the random variable. The dose of the baseline visit was set to 0, while the dose of treatment visits one and two were set to 1 and 2 respectively.

*Task data.* The pairwise differences between V1 and the treatment visits (V2 and V3) were tested for the fear-happy linear contrast using a linear mixed effects model. In this design, the fear-happy linear contrast Δβ value was the outcome variable, visit was the categorical variable of interest, the visit-specific negative valence score was an additional covariate, and patient was the random variable.

The pairwise differences between the happy-neutral and fear-neutral linear contrasts were tested for each treatment status (‘treatment’ or ‘baseline’) using paired two-sample *t*-tests. In these tests, the two groups were the two contrasts (happy-neutral and fear-neutral).

*Arousal and valence ratings.* The pairwise differences between the baseline visit and the treatment visits were tested for the arousal, negative valence, and positive valence scores using three linear mixed effects models. In these designs, arousal or valence scores were the outcome variable, visit was the categorical variable of interest, and patient was the random variable. The effect of iTBS dose on these scores was tested using three linear mixed effects models. In these designs, arousal or valence scores were the outcome variable, dose was the continuous covariate of interest, and patient was the random variable. The dose of the baseline visit was set to 0, while the dose of treatment visits one and two were set to 1 and 2 respectively.

*Simplified- Functional Movement Disorders Rating Scale (S-FMDRS) scores.* The difference between pre-iTBS S-FMDRS score and post-iTBS score was tested using a linear mixed effect model. In this design, the S-FMDRS score was the outcome variable, within visit time (pre- or post-iTBS) was the categorical variable of interest, and patient was the random variable. Only data from V2 and V3 were used to fit this model. *Post-hoc* paired two-sample *t*-tests were performed to compare pre-iTBS S-FMDRS score to post-iTBS S-FMDRS score within each visit (V2 and V3) separately.

The pairwise differences between the pre-iTBS S-FMDRS collected at V2 and the scores collected at pre-iTBS V3 and V4 were evaluated using a mixed linear effects model. In this design, the S-FMDRS score was the outcome variable, visit was the categorical covariate of interest, and patient was the random variable.

The difference between the S-FMDRS score collected at V1 and the score collected at pre-iTBS V2 was evaluated using a paired two-sample *t*-test.

Supplementary Figure Legends

Supplementary Figure 1. **Exploratory imaging analysis results.** Panel A shows changes in rsFC between the individualized DLPFC target and the right amygdala. The y axis represents changes in z-transformed region-to-region correlation strength as a result of iTBS. Panel B shows changes in right amygdala reactivity to fearful to neutral (F-N) vs. happy – neutral (H-N) faces during the fMRI task. The y axis represents left amygdala beta values. Error bars represent SEM.

Supplementary Figure 1.

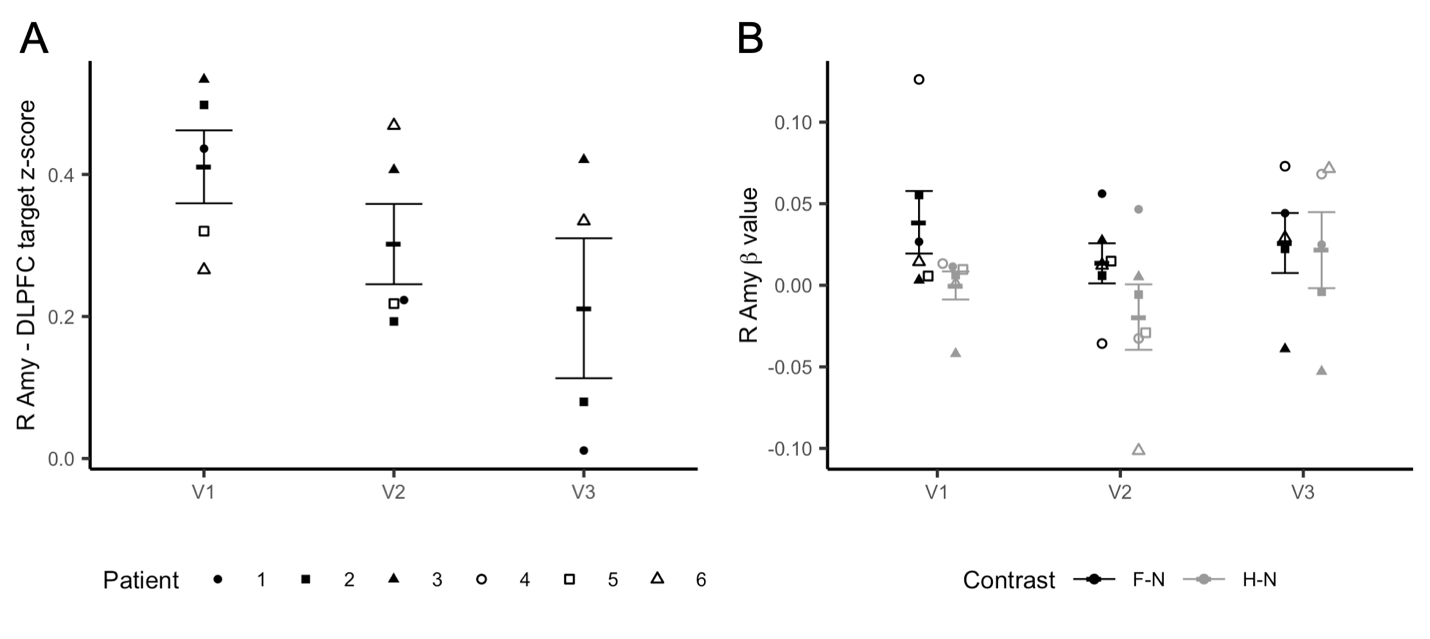
